# INCLUSIVE HEALTH: MODELING COVID-19 IN CORRECTIONAL FACILITIES AND COMMUNITIES

**DOI:** 10.1101/2021.02.05.21251174

**Authors:** Scott Greenhalgh, Ashley Provencher

**Affiliations:** Siena College, Department of Mathematics, 515 Loudon Road, Loudonville, NY, 12211, USA.; Siena College, Department of Economics, 515 Loudon Road, Loudonville, NY, 12211, USA.

**Keywords:** Mathematical model, Corrections, COVID-19, Incarceration

## Abstract

Mass incarceration, commonly associated with overcrowding and inadequate health resources for incarcerated people, creates a fertile environment for the spread of the coronavirus disease 2019 (COVID-19) in U.S. correctional facilities. The exact role that correctional facilities play in enhancing COVID-19 spread and enabling community re-emergence of COVID-19 is unknown. We constructed a novel stochastic model of COVID-19 transmission to estimate the impact of correctional facilities, specifically jails and state prisons, for enhancing disease transmission and enabling disease re-emergence in local communities. Using our model, we evaluated scenarios of testing and quarantining infected incarcerated people at 0.0, 0.5, and 1.0 times the rate that occurs for infected people in the local community for population sizes of 5, 10, and 20 thousand people. Our results illustrate testing and quarantining an incarcerated population of 800 would reduce the probability of a major community outbreak by 6% and also prevent between 250 to 730 incidences of COVID-19 per year, depending on local community size. These findings illustrate that managing COVID-19 in correctional facilities is essential to mitigate risks to community health, and thereby stresses the importance of improving the health standards of incarcerated people.

---

Cramped and overpopulated, correctional facilities are ideal environments for viruses to spread. This was made clear with the ongoing rapid spread of the coronavirus disease 2019 (COVID-19) in US jails and prisons. As US jails and prisons are structurally designed for communal living to efficiently confine people, the rate of infection is 5.5 times higher in U.S. state and federal prisons than in the broader community (1). Limited access to personal protective equipment, hand sanitizer, and even soap exacerbates spread across all people within the facility. In prisons, 366,121 incarcerated people and 98,035 correctional staff have been diagnosed with COVID-19 as of January 2021 (2).

In contrast with other group living quarters, such as nursing facilities, correctional facilities enable disease persistence and re-emergence. The reason for this is that many of the people who are housed within a correctional facility do not stay there for a long time. In the United States, on average, a person is confined to jail for 25 days (3), a state prison for 2.6 years (4), or to federal prison for 4.5 years (5). Correctional facilities, particularly jails, thus serve as reservoirs that enable disease persistence because people continually enter them without prior exposure to the virus, which facilitates its spread. Likewise, regular exits from correctional facilities may result in the virus’ reemergence within the local community if its spread is unchecked.

It is unknown at what level correctional facilities contribute to the persistence of COVID-19 in the local community or how the implementation of COVID-19 control measures might mitigate these risks. So, to inform on these issues, we construct a new stochastic model of COVID-19 transmission to estimate the impact of correctional facilities for enhancing disease transmission and enabling disease re-emergence. We conclude with a discussion of potential strategies to prevent disease persistence and re-emergence between correctional facilities and their local communities.

## METHODS

We constructed a novel stochastic model of COVID-19 transmission to estimate the impact of correctional facilities in enabling disease transmission and disease re-emergence. We calibrate our model to describe COVID-19 spread in communities with correctional facilities that house 800 incarcerated people and staffed with 420 correctional workers. For such a facility, we measure how testing and quarantining infected incarcerated people at 0.0, 0.5, 1.0 times the rate that occurs for the general population impacts the spread of COVID-19 within the correctional facility and among the broader community. In addition, to reflect the different population densities of communities near correctional facilities, we also consider small, medium, and large communities of 5,000, 10,000, and 20,000 people, respectively, in addition to the average incarceration period of 25 days in jails and 2.6 years in state prisons.

### Mathematical model

In the stochastic model, we consider a population segregated as the local community (C), incarcerated people (P), and correctional workers (W), where correctional workers are defined as civilian employees or volunteers who reside outside of the correctional facility. We further subdivide each section of the population based on COVID-19 infection status using subscripts to denote susceptible individuals (S), latently infected individuals (E), asymptomatic infected individuals (A), symptomatic infected individuals (I), individuals that have recovered from infection (R), and individuals that test positive for COVID-19 infection and are quarantined (Q).

The rate at which susceptible individuals in the local community acquire COVID-19 is given by the force of infection:

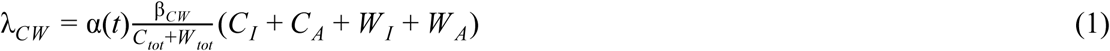

where β_*CW*_ is the transmission rate of COVID-19 in the local community, *C*_*tot*_ is the total size of the community, *W*_*tot*_ is the total size of the correctional workers, and α(*t*) is the impact of public health control measures, such as face masks and social distancing, on mitigating COVID-19 spread in the local community. Similarly, the rate at which susceptible correctional workers acquire COVID-19 is given by the force of infection:

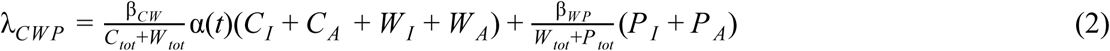

where β_*WP*_ is the transmission rate of COVID-19 in the correctional workers. Finally, the rate at which susceptible incarcerated people acquire COVID-19 is given by the force of infection:

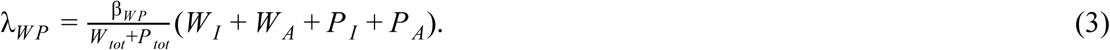

To reflect the influence of social distancing efforts on the transmission of COVID-19 in the local community, we consider the distinct phases of 1) pre-social distancing and 2) social distancing, along with the gradual fade-out of social distancing (Web Appendix 1, Web Figure 2). These phases are reflected in the rate new infections occur through the function

**Figure 1.**
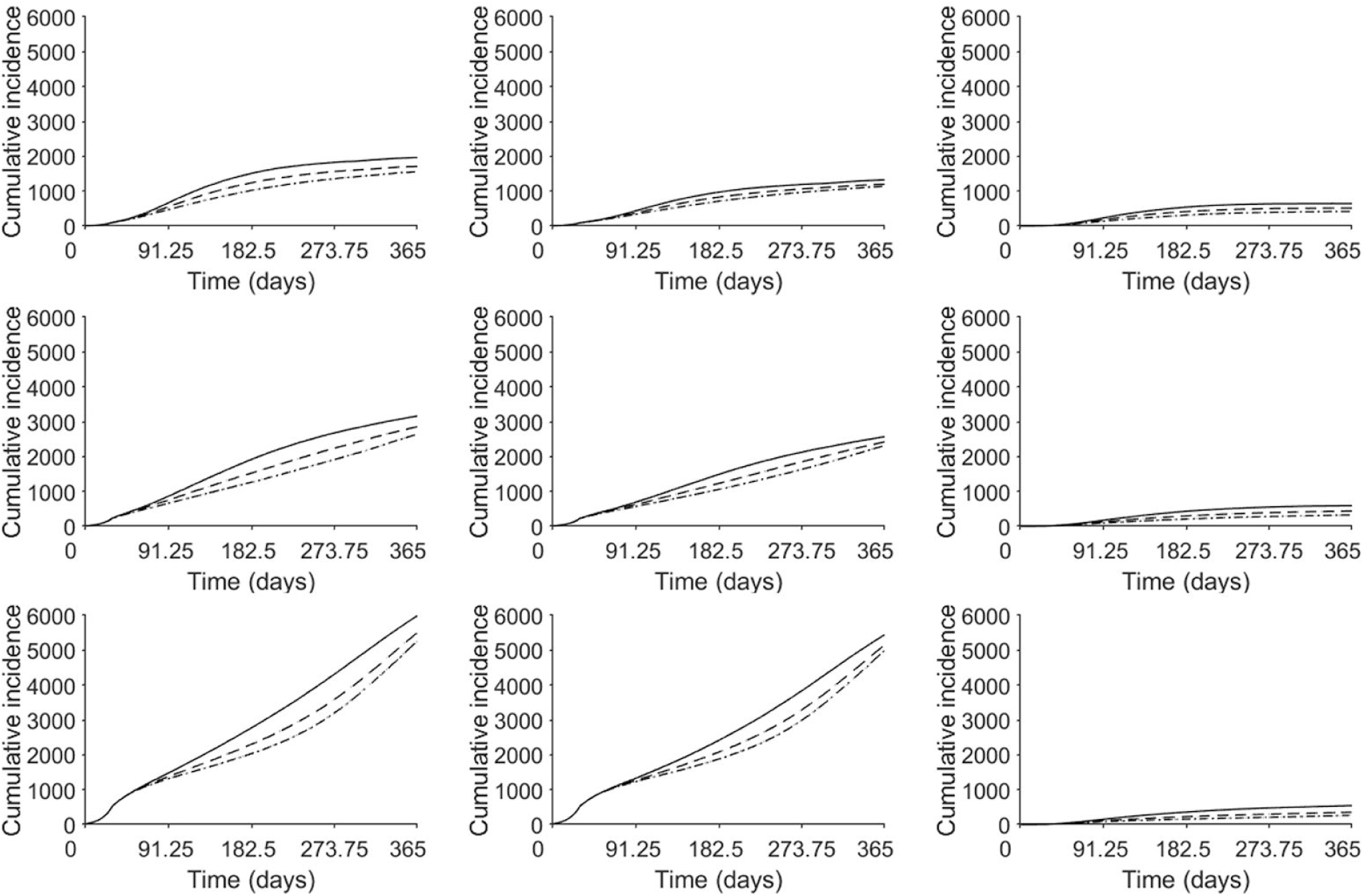
Cumulative COVID-19 incidence. Predicted COVID-19 incidence in the entire population (top), the local community (middle), and in the correctional facility (bottom) for population sizes of 5,000 people (left), 10,000 people (center), and 20,000 people (right). Predicted COVID-19 incidence is illustrated for no intervention in correctional facilities (solid black line), an intervention where testing and quarantining occur at 0.5 times the rate of the general population (dashed black line), and 1.0 times the rate of the general population (dotted black line) where the average duration of incarceration is 25 days.

**Figure 2.**
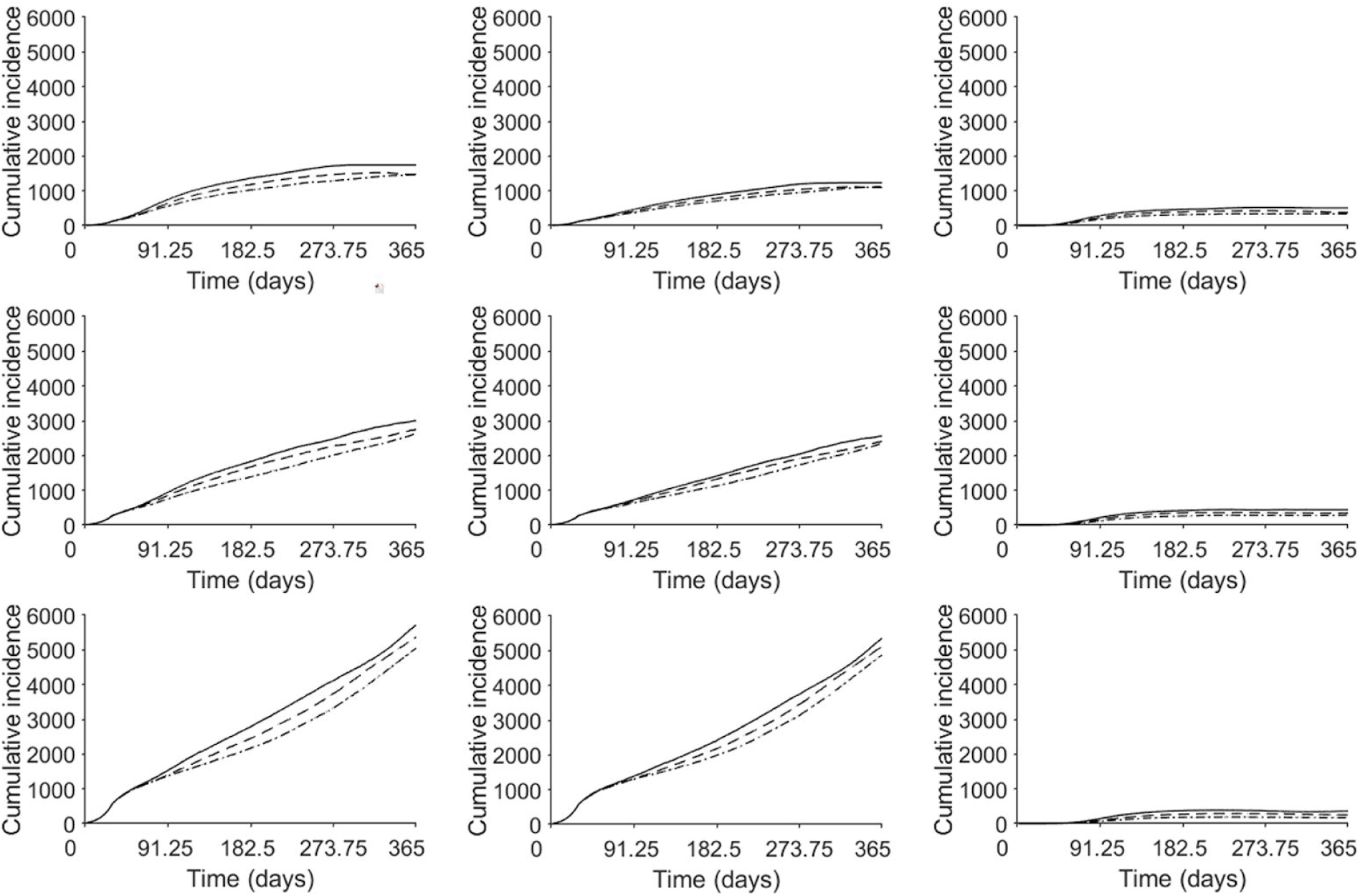
Cumulative COVID-19 incidence. Predicted COVID-19 incidence in the entire population (top), the local community (middle), and in the correctional facility (bottom) for population sizes of 5,000 people (left), 10,000 people (center), and 20,000 people (right). Predicted COVID-19 incidence is illustrated for no intervention in correctional facilities (solid black line), an intervention where testing and quarantining occur at 0.5 times the rate of the general population (dashed black line), and 1.0 times the rate of the general population (dotted black line) where the average duration of incarceration is 2.6 years.

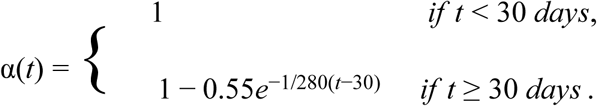

Further details and additional model parameters, including the COVID-19 latent period, infectious period, the proportion of COVID-19 asymptomatic infections, are available in Table 1.

**Table 1.**
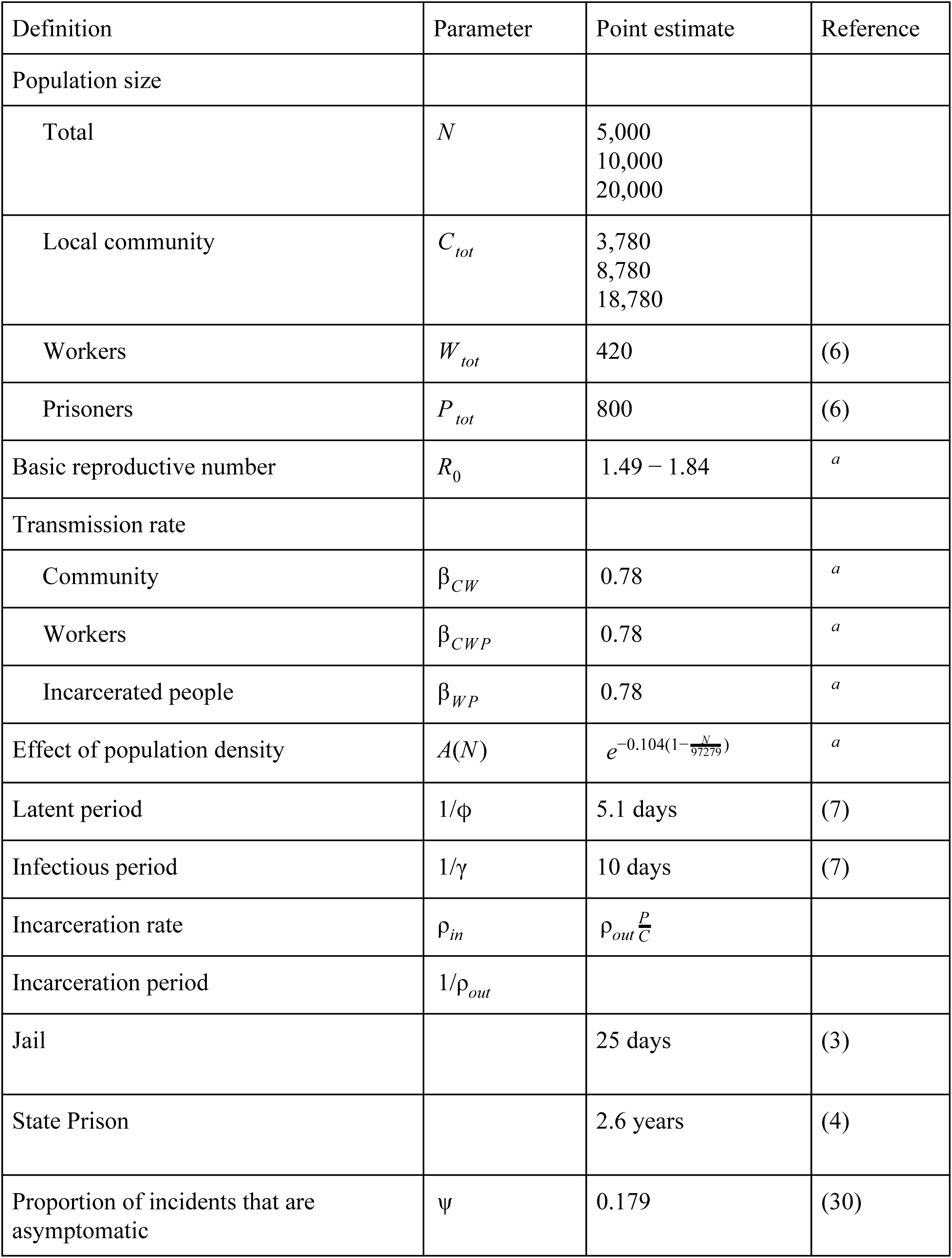
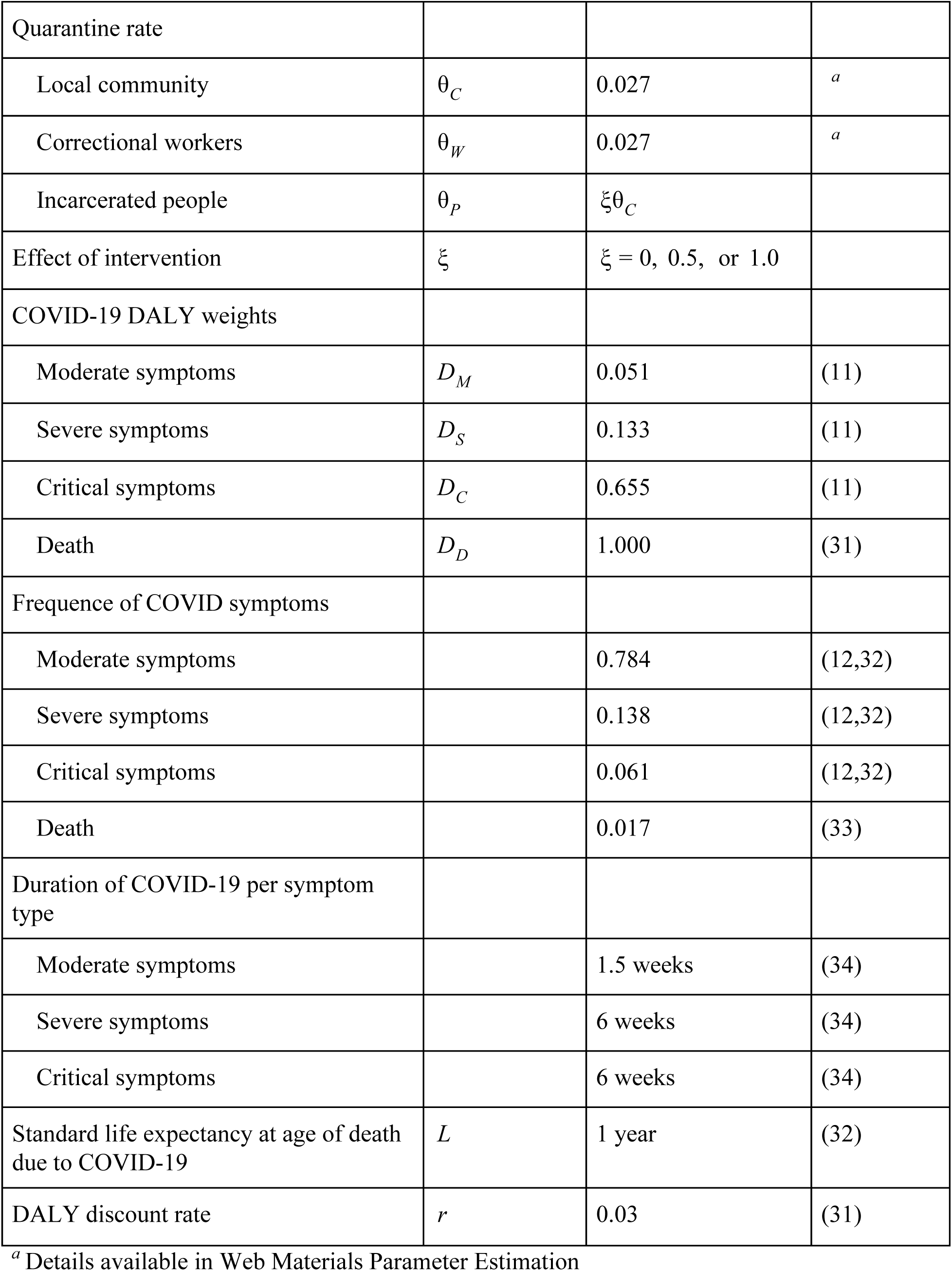
Parameter names, values, and sources.

| Definition | Parameter | Point estimate | Reference |
| --- | --- | --- | --- |
| Population size |  |  |  |
| Total | $N$ | 5,000<br>10,000<br>20,000 | |
| Local community | $C_{tot}$ | 3,780<br>8,780<br>18,780 | |
| Workers | $W_{tot}$ | 420 | (6) |
| Prisoners | $P_{tot}$ | 800 | (6) |
| Basic reproductive number | $R_0$ | 1.49 – 1.84 | <sup>a</sup> |
| Transmission rate |  |  |  |
| Community | $\beta_{CW}$ | 0.78 | <sup>a</sup> |
| Workers | $\beta_{CWP}$ | 0.78 | <sup>a</sup> |
| Incarcerated people | $\beta_{WP}$ | 0.78 | <sup>a</sup> |
| Effect of population density | $A(N)$ | $e^{-0.104(1-\frac{N}{97279})}$ | <sup>a</sup> |
| Latent period | $1/\phi$ | 5.1 days | (7) |
| Infectious period | $1/\gamma$ | 10 days | (7) |
| Incarceration rate | $\rho_{in}$ | $\rho_{out} \frac{P}{C}$ | |
| Incarceration period | $1/\rho_{out}$ | | |
| Jail |  | 25 days | (3) |
| State Prison |  | 2.6 years | (4) |
| Proportion of incidents that are asymptomatic | $\psi$ | 0.179 | (30) |
| Quarantine rate |  |  |  |
| Local community | $\theta_C$ | 0.027 | <sup>a</sup> |
| Correctional workers | $\theta_W$ | 0.027 | <sup>a</sup> |
| Incarcerated people | $\theta_P$ | $\xi\theta_C$ | |
| Effect of intervention | $\xi$ | $\xi = 0, 0.5, \text{ or } 1.0$ | |
| COVID-19 DALY weights |  |  |  |
| Moderate symptoms | $D_M$ | 0.051 | (11) |
| Severe symptoms | $D_S$ | 0.133 | (11) |
| Critical symptoms | $D_C$ | 0.655 | (11) |
| Death | $D_D$ | 1.000 | (31) |
| Frequency of COVID symptoms |  |  |  |
| Moderate symptoms |  | 0.784 | (12,32) |
| Severe symptoms |  | 0.138 | (12,32) |
| Critical symptoms |  | 0.061 | (12,32) |
| Death |  | 0.017 | (33) |
| Duration of COVID-19 per symptom type |  |  |  |
| Moderate symptoms |  | 1.5 weeks | (34) |
| Severe symptoms |  | 6 weeks | (34) |
| Critical symptoms |  | 6 weeks | (34) |
| Standard life expectancy at age of death due to COVID-19 | $L$ | 1 year | (32) |
| DALY discount rate | $r$ | 0.03 | (31) |

<sup>a</sup> Details available in Web Materials Parameter Estimation

### Parameter estimation

Model population sizes for the number of incarcerated people and correctional workers were determined using publicly available data on the capacity and annual occupancy of correctional facilities (6). The model also uses current estimates of the duration of infection and the latent period of COVID-19 from the literature (7). In addition, the impact of pre- and post-social distancing (8,9) on COVID-19 transmission in the local community was incorporated into model transmission rates (Web Appendix 1, Web Figure 2). Specifically, to estimate model transmission rates, we fit a deterministic analog of our stochastic model (Web Tables 1-4) to COVID-19 incidence, use the estimated ratio of undetected and detected cases of 4.64:1 in New York state, in addition to New York state data on the number of positive COVID-19 incidents (8,9). We also determined the rate of testing and quarantining of infected people from the local community (Web Appendix 2, Web Figure 3) using available data on positive COVID-19 tests (8).

**Figure 3.**
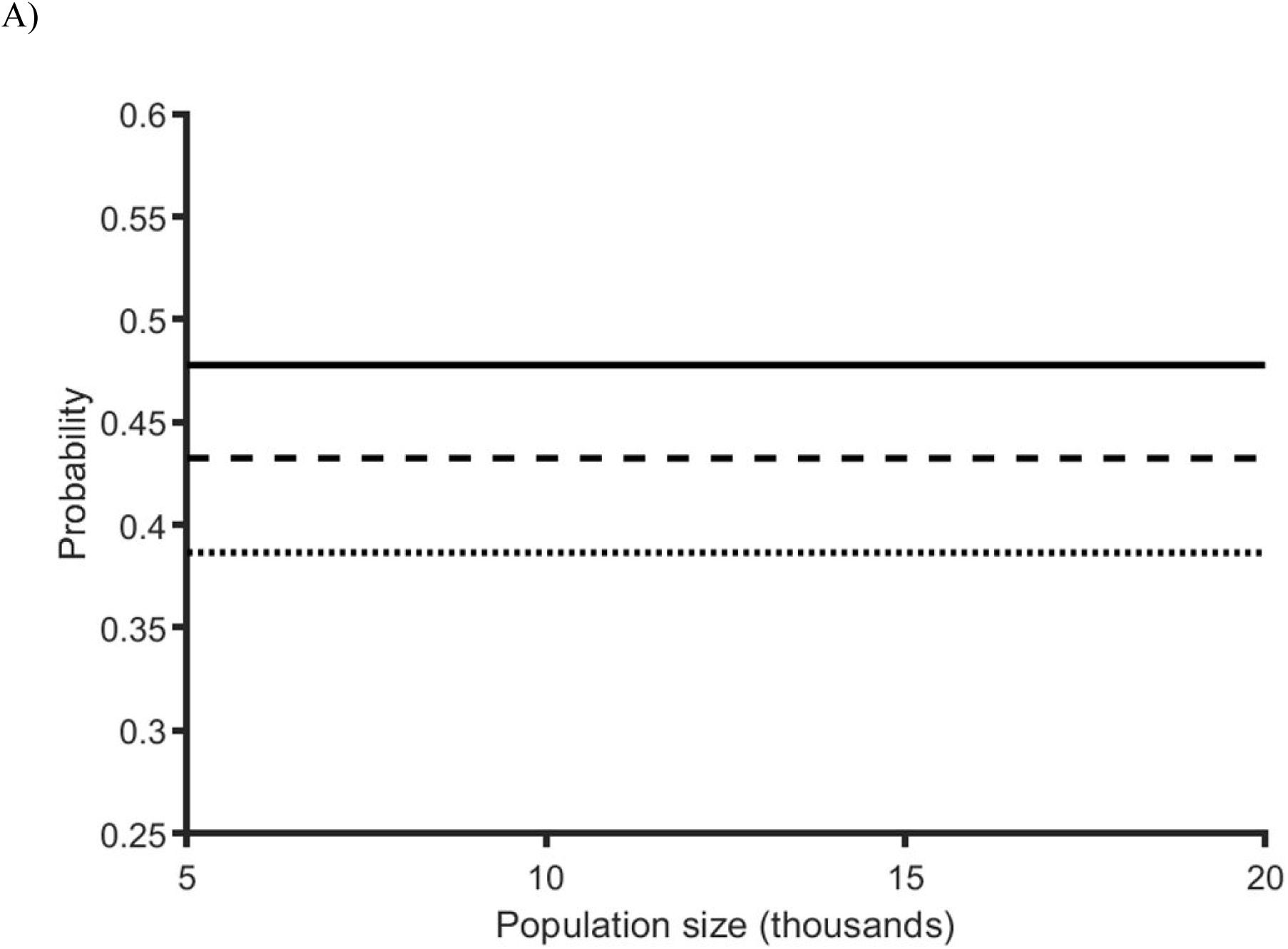
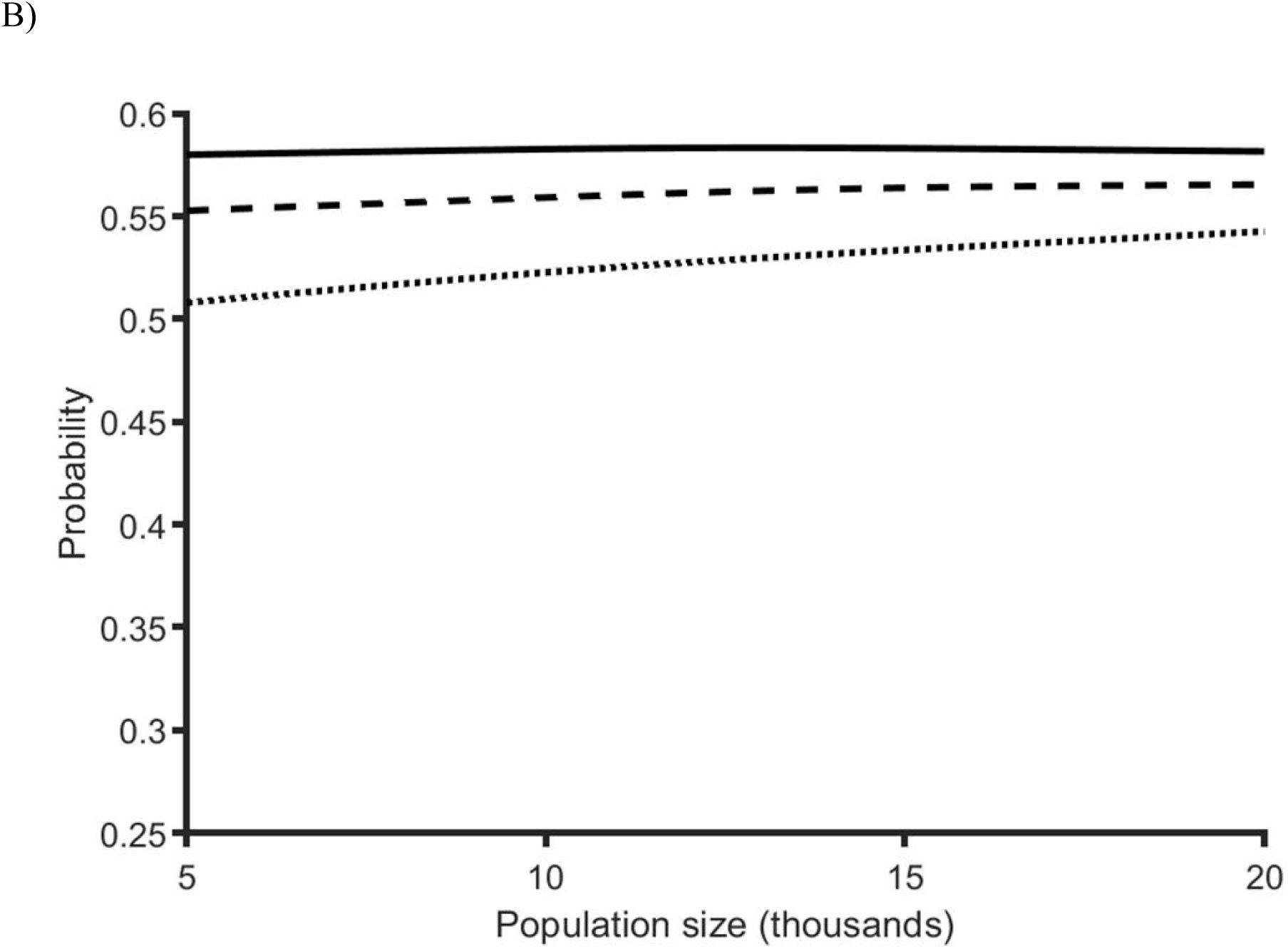
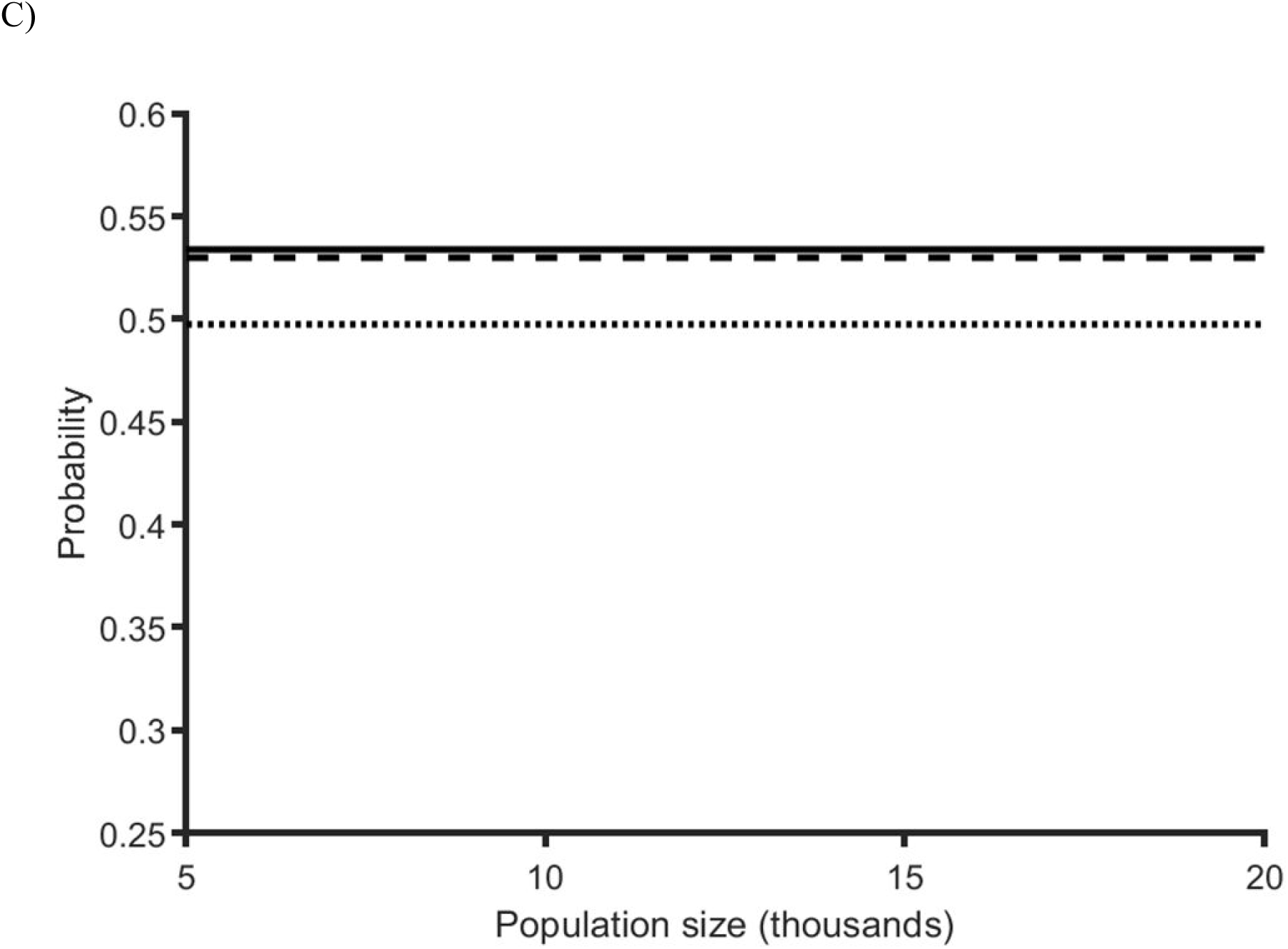
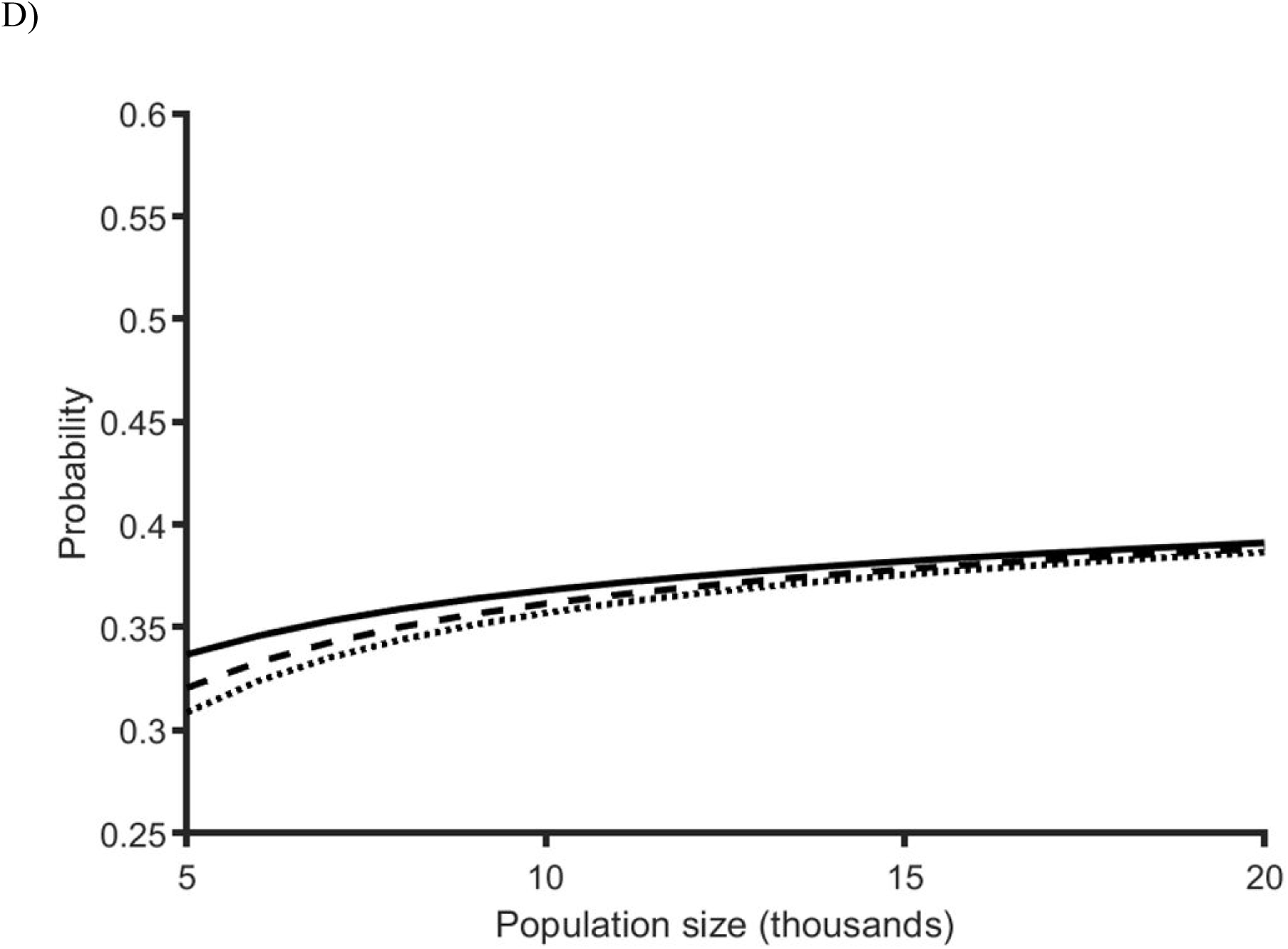
Probability of major outbreaks and incarceration-to-community transmission. The probability that a single COVID-19 infected incarcerated person results in A) a major outbreak in a correctional facility, B) a community transmission event in a jail when no quarantining and testing of incarcerated people occurs (solid black line), when testing occurs at 0.5 the rate of the local community (dashed black line) and when testing occurs at the same rate as the local community (dotted black line), C) a community transmission event in a state prison when no quarantining and testing of incarcerated people occurs (solid black line), when testing occurs at 0.5 the rate of the local community (dashed black line) and when testing occurs at the same rate as the local community (dotted black line), D) The probability that a single COVID-19 infected person results in a major outbreak in the local community when no quarantining and testing of incarcerated occurs (solid black line), when testing occurs at 0.5 the rate of the local community (dashed black line) and when testing occurs at the same rate as the local community (dotted black line).

### Intervention scenarios

To inform on the effect of identifying COVID-19 infected incarcerated people on transmission, we consider scenarios where the testing and quarantining rate of incarcerated people is 0, 0.5, and 1.0 times the rate used in the local community. We consider these testing and quarantining rates for total population sizes of 5,000, 10,000, and 20,000 people, where the rate of incarceration and average duration of incarceration reflect those occurring in jails and state prisons.

### Health outcomes

To determine the direct and indirect benefit of identifying and quarantining COVID-19 infected people on death and disability due to COVID-19, we measure health outcomes in incidence averted and disability adjusted life-years (DALYs) (10). Specifically, we calculate time-discounted DALYs lost to COVID-19 over 1 year and determine the net DALYs saved by subtracting the total DALYs lost under no testing and quarantining from scenarios that consider identifying and quarantining COVID-19 infected incarcerated people. Disability weights for the DALY calculation and the proportion of people that endure each severity of COVID-19 are obtained from the literature (11–13).

### Community re-emergence

To provide insight on the potential role of correctional facilities in enabling COVID-19 reemergence, we estimate the likelihood of a correctional facility re-introducing COVID-19 into the local community. Specifically, we estimate the probability of a major outbreak (14) within the correctional facility and the local community for total population sizes of 5,000, 10,000, and 20,000 people. In addition, to provide insight on the risk of transmission from the correctional facility to the local community, we determine the proportion of 10,000 simulations where a single infection in the correctional facility leads to a transmission event in the local community.

## RESULTS

To inform on the potential role that correctional facilities play in COVID-19 transmission and the effect of testing and quarantining infected incarcerated people, we simulated transmission among incarcerated people, correctional workers, and people in the local community. Specifically, we evaluated scenarios where infected incarcerated people are identified at rates that are 0, 0.5, and 1.0 times that of the rate of the local community, with community population sizes of 5,000, 10,000, and 20,000 people. Our findings illustrate that testing and quarantining infected incarcerated people substantially reduced COVID-19 incidence and saved DALYs (Table 2, Table 3). Our findings also illustrate that smaller community sizes receive a greater reduction in COVID-19 incidence from testing and quarantining infected incarcerated people, in addition to a greater decrease in the risk for COVID-19 re-emergence.

**Table 2.** Base annual COVID-19 incidence, annual incidence averted/1,000, and annual DALYs saved/1,000 People when the average duration of incarceration is 25 days.

| | $N = 5,000$ | $N = 10,000$ | $N = 20,000$ |
| --- | --- | --- | --- |
| No intervention ($\theta_P = 0.0 \theta_C$) | | | |
| Baseline annual incidence (1,000s) | 1.96 | 3.16 | 5.98 |
| Baseline annual incidence for incarcerated people (1000's) | 0.64 | 0.59 | 0.54 |
| Baseline total DALYs per 1,000 people | 56.05 | 28.03 | 14.01 |
| 50% quarantine rate ($\theta_P = 0.5 \theta_C$) | | | |
| Annual incidence averted (1,000's) |  |  |  |
| Total population | 0.26 | 0.31 | 0.49 |
| Local community | 0.12 | 0.15 | 0.30 |
| Incarcerated population | 0.13 | 0.16 | 0.19 |
| Total DALYs saved per 1000 people | 9.55 | 4.32 | 2.01 |
| 100% quarantine rate ($\theta_P = 1.0 \theta_C$) | | | |
| Annual incidence averted (1,000's) |  |  |  |
| Total population | 0.41 | 0.52 | 0.73 |
| Local community | 0.18 | 0.26 | 0.45 |
| Incarcerated population | 0.23 | 0.26 | 0.28 |
| Total DALYs saved per 1,000 people | 15.31 | 8.56 | 3.01 |

**Table 3.**
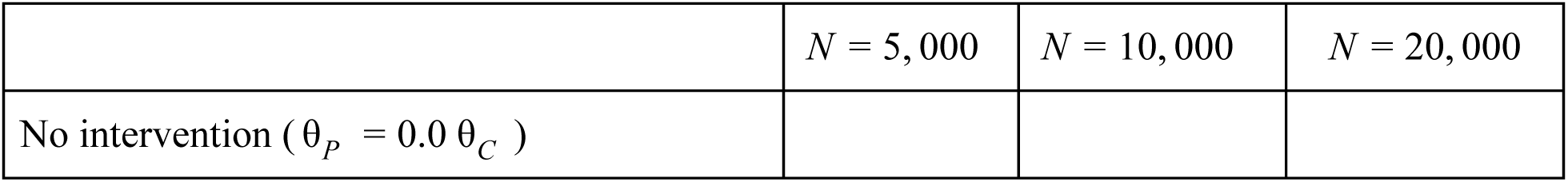
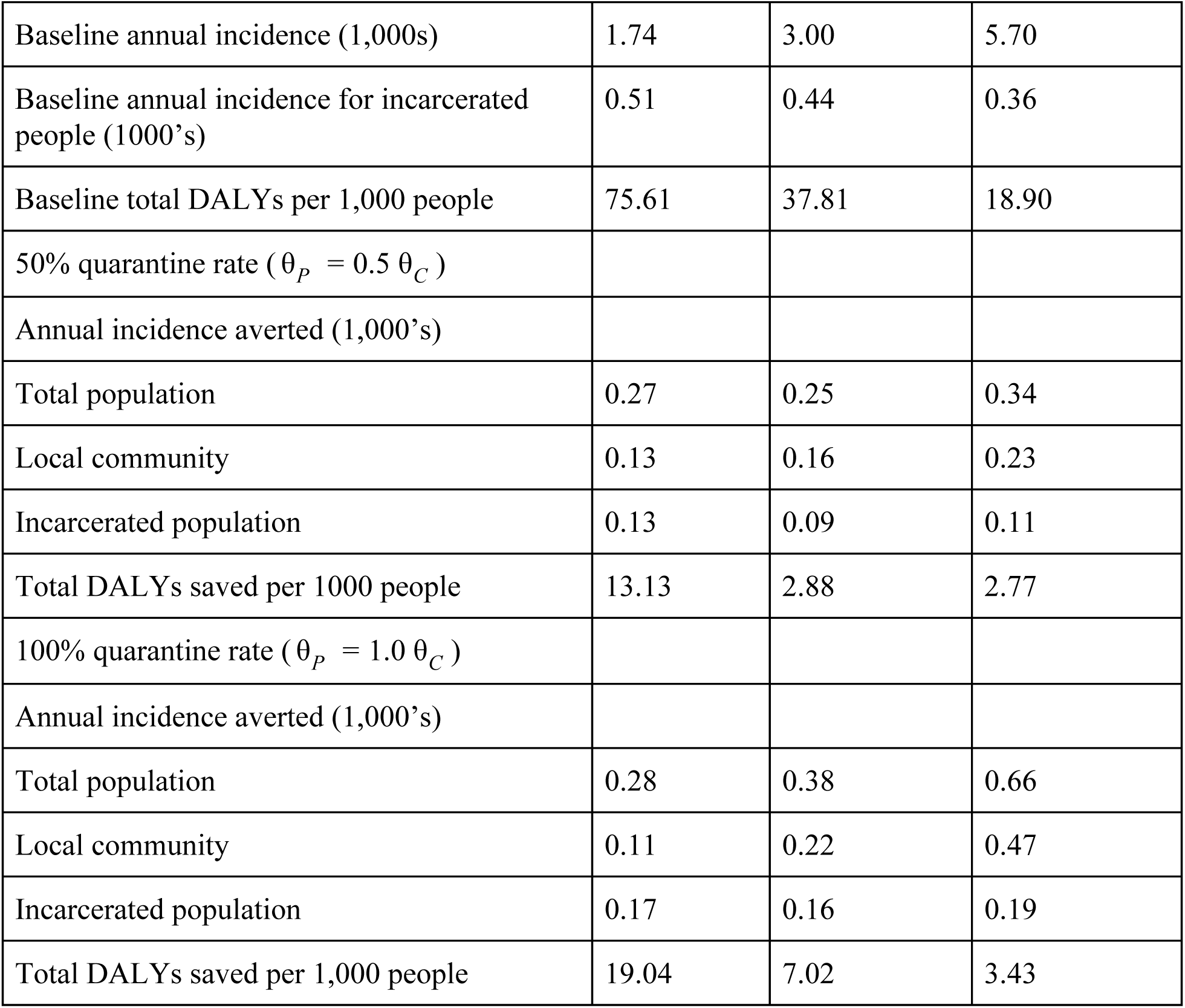
Base annual COVID-19 incidence, annual incidence averted/1,000, and annual DALYs saved/1,000 People when the average duration of incarceration is 2.6 years.

| | $N = 5,000$ | $N = 10,000$ | $N = 20,000$ |
| --- | --- | --- | --- |
| No intervention ($\theta_P = 0.0 \theta_C$) | | | |
| INCLUSIVE HEALTH |  |  |  |
| Baseline annual incidence (1,000s) | 1.74 | 3.00 | 5.70 |
| Baseline annual incidence for incarcerated people (1000's) | 0.51 | 0.44 | 0.36 |
| Baseline total DALYs per 1,000 people | 75.61 | 37.81 | 18.90 |
| 50% quarantine rate ( $\theta_P = 0.5 \theta_C$ ) | | | |
| Annual incidence averted (1,000's) |  |  |  |
| Total population | 0.27 | 0.25 | 0.34 |
| Local community | 0.13 | 0.16 | 0.23 |
| Incarcerated population | 0.13 | 0.09 | 0.11 |
| Total DALYs saved per 1000 people | 13.13 | 2.88 | 2.77 |
| 100% quarantine rate ( $\theta_P = 1.0 \theta_C$ ) | | | |
| Annual incidence averted (1,000's) |  |  |  |
| Total population | 0.28 | 0.38 | 0.66 |
| Local community | 0.11 | 0.22 | 0.47 |
| Incarcerated population | 0.17 | 0.16 | 0.19 |
| Total DALYs saved per 1,000 people | 19.04 | 7.02 | 3.43 |

A single infected incarcerated person is more likely to lead to a community transmission event than causing a major COVID-19 outbreak within the correctional facility (Figure 3A, Figure 3B, Figure 3C). Specifically, the probability that a single infected incarcerated person leads to a major outbreak of COVID-19 within a facility is 0.48, 0.43, or 0.39 (Figure 3A), whereas the probability a single infected incarcerated person in jail causes a community transmission event is 0.56, 0.52, and 0.50 (Figure 3B), depending on whether quarantining and testing occurs at 0.0, 0.5, or 1.0 times the rate of the local community, respectively. Furthermore, while the probability that a single infected incarcerated person in a state prison causes a community transmission event is lower than that of a jail, with probabilities of 0.53, 0.53, and 0.50 (Figure 3C), respectively, they remain higher than the probability of a COVID-19 outbreak in the correctional facility. Upon the successful transmission of COVID-19 to outside the correctional facility, we found the probability of a major outbreak of COVID-19 in the local community increases with the population (Figure 3D). For a population size of 20,000 people, testing and quarantining had a negligible impact on this probability, with the probability of a major outbreak for all scenarios of 0.46 approximately. However, for a population size of 5,000 people, testing and quarantining incarcerated people decreased the probability of a major outbreak from 0.48 to 0.43 when testing and quarantining occur at 0.5 the level of the local community, and to 0.39 when testing and quarantining occur at the same rate as the local community.

When considering a total population size of 5,000 people that feature a nearby jail, we predicted 1.96 annual incidents of COVID-19 per 1,000 people annually, with 1.32 of these incidents occurring in the local community, and 0.64 occurring within the correctional facility (Table 2). Through testing and quarantining infected incarcerated people, 260 and 410 annual incidents of COVID-19 can be averted when correctional facilities test and quarantine at 0.5 and 1.0 times the rate of the local community, respectively (Table 2). Our results also illustrate the local community benefits nearly as much as the incarcerated people from their testing and quarantining, concerning the reduction in COVID-19 incidence (Table 2), and annually saves 15.31 to 75.61 DALYs per 1,000 people depending on the testing and quarantine scenario.

For a total population size of 10,000 people, COVID-19 incidence increased to 3.16 annual incidents per 1,000 people, with 2.57 and 0.59 annual incidents per 1,000 people occurring within the local community and correctional facility, respectively (Table 2). Through testing and quarantining infected incarcerated people at 0.5 and 1.0 times the rate of the local community, 3,100 and 520 annual incidents of COVID-19 can be averted, respectively, with nearly half of the averted incidents occurring in the local community (Table 2). In addition, testing and quarantining incarcerated people at 0.5 the rate of the general public annually saves 4.32 DALYs per 1,000 people, with this number increases to 8.56 DALYs per 1,000 people when testing and quarantining occurred at the same rate.

With a total population size of 20,000 people, COVID-19 incidence occurred at 5.98 annual incidents per 1,000 people. Of these incidents, 5.44 and 0.54 incidents per 1,000 people occur in the local community, and the correctional facility, respectively (Table 2). By testing and quarantining infected incarcerated people at 0.5 and 1.0 times the rate of the local community, we found that 490 and 730 annual incidents of COVID-19 can be averted (Table 2).

Furthermore, from these total incidents averted, the majority occurs in the local community with 300 to 450 annual incidents of COVID-19 being averted, depending on the testing and quarantining rate. In addition, testing and quarantining incarcerated people at 0.5 the rate of the general public annually saves 2.01 DALYs per 1,000 people, with this number increasing to 3.01 DALYs per 1,000 people when testing and quarantining occurred at the same rate.

For COVID-19 transmission in state prisons, our model predicts 1.74, 3.00, and 5.70 annual incidents per 1,000 people for total population sizes of 5,000, 10,000, and 20,000 people, respectively. Through testing and quarantining at state prisons, these numbers can be reduced by 270 to 280, 250 to 380, and 340 to 660 incidences, respectively, depending on whether testing and quarantining incarcerated people occurs at 0.5 or 1.0 times the rate of the local population. To elaborate, if testing and quarantining incarcerated people occurs at 0.5 or 1.0 times the rate of the local population then 110 to 470 incidences of COVID-19 are averted in the local community, and 90 to 190 incidences of COVID-19 are averted in the state prison, depending on total population size (Table 3).

## DISCUSSION

The analysis of our model of COVID-19 transmission between correctional facilities and local communities illustrates that testing and quarantining incarcerated people substantially reduces the health burden of COVID-19. Specifically, our model’s predictions illustrate that testing and quarantining incarcerated people reduces COVID-19 incidence in both correctional facilities and local communities, reduces the likelihood of outbreaks, and saves DALYs in both jails and state prisons.

Our work highlights a critical public health challenge: COVID-19 persists within correctional facilities and these facilities are likely to reintroduce the virus into local communities. At the forefront of what enables this public health challenge is that correctional facilities offer a reservoir of susceptible people that constantly changes given their short duration of incarceration.

While jails and state prisons have different average durations of incarceration both are not closed systems that operate exclusively outside of local communities. Local communities experiencing an outbreak of COVID-19 will likely lead to an outbreak within such correctional facilities and *vice versa*. Highlighting this connection is the experience of Cook County where nearly 16% of COVID-19 incidents were traced to the local jail (15). Indeed, our simulations corroborate that the fallout from an outbreak within a correctional facility is dire for everyone. Fortunately, according to our results, testing and quarantining incarcerated individuals will substantially reduce the health burden of COVID-19 in both correctional facilities and local communities. Specifically, testing and quarantining incarcerated people reduces the risks for major COVID-19 outbreaks and cross-transmission, causes a reduction in incidence in both correctional facilities and nearby communities, and saves DALYs. All together these reduced risks and reductions provide strong motivation for the adoption of health policy that explicitly includes the health of incarcerated people when addressing local community health.

Although our work illustrates a health benefit for testing and quarantining incarcerated individuals, a single policy is not sufficient to prevent outbreaks across all correctional facilities and local communities. To elaborate, in the early days of the pandemic, many policymakers quickly adopted quarantine and early release policies to achieve greater social distancing within correctional facilities (16). The populations of jails and prisons have declined by 20% and 5%, respectively (17). Of course, these policies are more difficult to enact in some facilities than others, which stresses that adopting one policy is not likely the most effective strategy to reduce virus spread. Furthermore, both the environment within the correctional facility and the local community are important when determining COVID-19 mitigation and prevention strategies. For instance, our results illustrate that in local communities with relatively small populations, the incidence of new cases stands to decline greatly if the correctional facility were able to quarantine incarcerated people or test incarcerated people at least at the same rate as people in the local community. In contrast, while testing and quarantining reduce COVID-19 incidence in larger populations, it is less effective for curtailing the outbreak. This finding for larger facilities and communities suggests policies that reduce the number of incarcerated people, such as early release, are needed to diminish correctional facilities’ capacity to act as superspreading environments.

Reducing the number of incarcerated people is one policy to mitigate the superspreading potential of incarceration facilities, but it is not the only one. With the development of COVID-19 vaccines, advocates and health policy researchers have called on policymakers to make vaccines available to incarcerated people during the earliest phases of distribution (27). While this policy would likely mitigate the superspreading potential of correctional facilities, not all states are adopting it, and the majority of those that have are prioritizing vaccine distribution to correctional staff and exclude incarcerated people (28). Such actions, according to our findings, illustrate a lost opportunity to maximize health and safety, and suggest a more inclusive approach to COVID-19 vaccination would benefit everyone.

Policies aiming to reduce outbreaks within correctional facilities and local communities should be health-informed. Social distancing practices such as changes to housing or severe lockdowns within cells may mitigate spread within facilities, but will likely harm the mental wellbeing of incarcerated people (18, 19). In contrast, issuing telephone cards for incarcerated people to stay in contact with family could improve mental wellbeing (20). Other policies that improve sanitation, including access to disinfectants and personal protective equipment (20), or improve access to quality healthcare for incarcerated people, such as greater use of telemedicine (21), mitigate virus spread in correctional settings.

Findings from this study are limited in a few ways. For instance, data on the contacts between people in correctional facilities and the local community are limited, although such limitations typically do not impede the widespread use of stochastic models in the study of disease transmission. We also do not account for the declining number of people in correctional facilities prior to the pandemic (22), nor the myriad decarceration policies that occurred once the pandemic was underway. Furthermore, with regards to the policies of testing and quarantining of infectious individuals, our model assumes that these occur simultaneously, and does not account for the fact that their separate combination, through actions such as contact tracing and targeting at-risk persons, would likely save even more lives and mitigate disease spread further.

Similarly, with regards to mitigating disease spread, our model assumes a standard population density in a correctional facility, whereas, not all correctional facilities have the same layout, particularly as it relates to housing for incarcerated people. While dormitories and cells are the most common types of housing units, the availability and use of these spaces can vary considerably across facilities. Others (23, 24) have found evidence that the type of facility housing matters, and that people housed in dormitories are more susceptible to contracting the virus. As such, future models would do well to incorporate information on the varied population densities within correctional housing spaces to better understand viral spread. Another important characteristic that future models should consider is the disproportional impact of the pandemic on minorities (25,26). To elaborate, Blacks and Hispanics are overrepresented by 5.6 and 3.0 times more than White adults (22) in U.S. correctional facilities, which contributes to disparities of these groups in COVID-19 testing, cases, and deaths. As such, while our results provide a uniform estimate on the benefits of quarantining and testing for these groups, future research is urgently needed that investigates the intersection of race, health, and criminal justice involvement to better understand how criminal justice policy and practice may exacerbate health disparities by race.

In summary, the health of incarcerated people likely has a substantial impact on the risk and magnitude of COVID-19 outbreaks in communities. Our findings illustrate that routine testing and quarantining of incarcerated people carries a dual benefit for correctional facilities, and their local communities. Thus, our work suggests that to maximize public health’s ability to combat the ongoing COVID-19 pandemic, incarcerated people’s well-being should be included in the design and implementation of health policies.

## Supporting information

Additional methods details

## Data Availability

All data is available within the manuscript

## Notes

### Competing Interest Statement

The authors have declared no competing interest.

### Funding Statement

No external funding was received.

