## Additional methods details for "INCLUSIVE HEALTH: MODELING COVID-19 IN CORRECTIONAL FACILITIES AND COMMUNITIES"

### Web Material

#### Table of Contents

Web Appendix 1: Transmission Rates

Web Appendix 2: Incarceration and Release Rates

Web Appendix 3: The Reproductive Numbers and the Probability of a Major Outbreak

Web Figure 1: Compartmental Diagram

Web Figure 2: Fit of model to COVID-19 data.

Web Figure 3: Fit of model to COVID-19 positive-test data.

Web Figure 4. Basic reproductive number for the local community.

Web Table 1: Transition probabilities for COVID-19 infection of incarcerated individuals

Web Table 2: Transition probabilities for COVID-19 infection of incarcerated workers

Web Table 3: Transition probabilities for COVID-19 infection of the general community

Web Table 4: Transition probabilities for the incarceration of general community members and the release of incarcerated individuals

#### Web Appendix 1: Transmission rates

**The COVID-19 transmission rate.** To estimate the transmission rate of COVID-19 for our stochastic model, we consider preliminary model based on a population divided into susceptible individuals ($S$), latently infected individuals ($E$), infected individuals ($I$), asymptomatic infected individuals ($A$), recovered individuals ($R$), and quarantined individuals ($Q$). Under this setup, we consider the force of infection given by

$$\lambda_{CW} = \frac{\beta_{CW}\alpha(t)(E+I)}{N}, \tag{S1}$$

where $N =$ and the transition between states of infection is given by,

$$\frac{dS}{dt} = -\lambda_{CW}S,$$

$$\frac{dE}{dt} = \lambda_{CW}S - \phi E - q_c E,$$

$$\frac{dI}{dt} = (1-\psi)\phi E - \gamma I - q_c I,$$

$$\frac{dA}{dt} = \psi\phi E - \gamma A - q_c A,$$

$$\frac{dR}{dt} = \gamma I + \gamma A,$$

$$\frac{dQ}{dt} = q_c(E + I + A).$$

Using this differential equation model, we estimate the transmission rate $\beta_{CW} \approx 0.78/day$ (Web Figure 2) using data on positive COVID-19 tests, the proportion of asymptomatic to symptomatic cases (1), and the ratio of reported to non-reported incidence (2).

**Social distancing function.** To reflect the effect of social distancing in the local community we fit a piecewise function to cummulative incidence data of COVID-19 in New York State (Web Figure 2). The peicewise function assumes that adherence to social distancing decays exponentially, and is given by

$$\alpha(t) = \begin{cases} 1 & \text{if } t < 30\ days, \\ 1 - 0.55e^{-\frac{1}{280}(t-30)} & \text{if t} \geq 30\ days. \end{cases}$$

#### Web Appendix 2: Incarceration and Release Rates

**COVID-19 testing and quarantine rate.** We estimate the testing and quarantining rate of the general community using available data on COVID-19 tests (3). Specifically, using a least squares procedure (Web Figure Y), we found that $\theta_C \approx 0.027/day$ (Web Figure 3).

**Incarceration rate.** To determine the rate of incarceration, $\rho_{in}$, we consider an incarceration facility with a capacity of 800 people that remain in the facility for an average duration of $1/\rho_{out}$ days. Therefore, for the facility to remain at capacity we require that inflow of newly incaracreated individuals equals the outflow of incarcerated individuals:

$$\rho_{in}\, C = \rho_{out} P,$$

where $C$ is the population size of the local community and $P$ is the  population size of the incarceration facility.

#### Web Appendix 3: The Reproductive Numbers and the Probability of a Major Outbreak

**The basic reproductive number.** To compute the basic reproductive number for COVID-19 infection we use standard next-generation techniques (4). Specifically, we use the stochastic model (Web Table 1-4), which has six compartments that contribute to transmission:  $C_I, C_A, W_I, W_A, P_I$, and $P_A$, along with routines for robust numerical calculation of Jacobian matrices (5). It follows that the basic reproductive number for within incarceration facilities is

$$R_0^P = \rho\left(F|_{\substack{DFE \\ \beta_{CW}=0}} V^{-1}\right),$$

and the basic reproductive number for local communities (Web Figure 4) is

$$R_0^C = \rho\left(F|_{\substack{DFE \\ \beta_{WP}=0}} V^{-1}\right),$$

where DFE stands for the disease-free equilibrium, and $\rho$ is the spectral radius operator.

**The probability of a major outbreak.** Given the definition of the basic reproductive numbers, the probability of a major outbreak in the incarceration facility given a single infected incarcerated individual (6) is approximately

$$1 - \frac{1}{R_0^P}.$$

Similarly, the probability of a major outbreak in the local community is

$$1 - \frac{1}{R_0^C}.$$

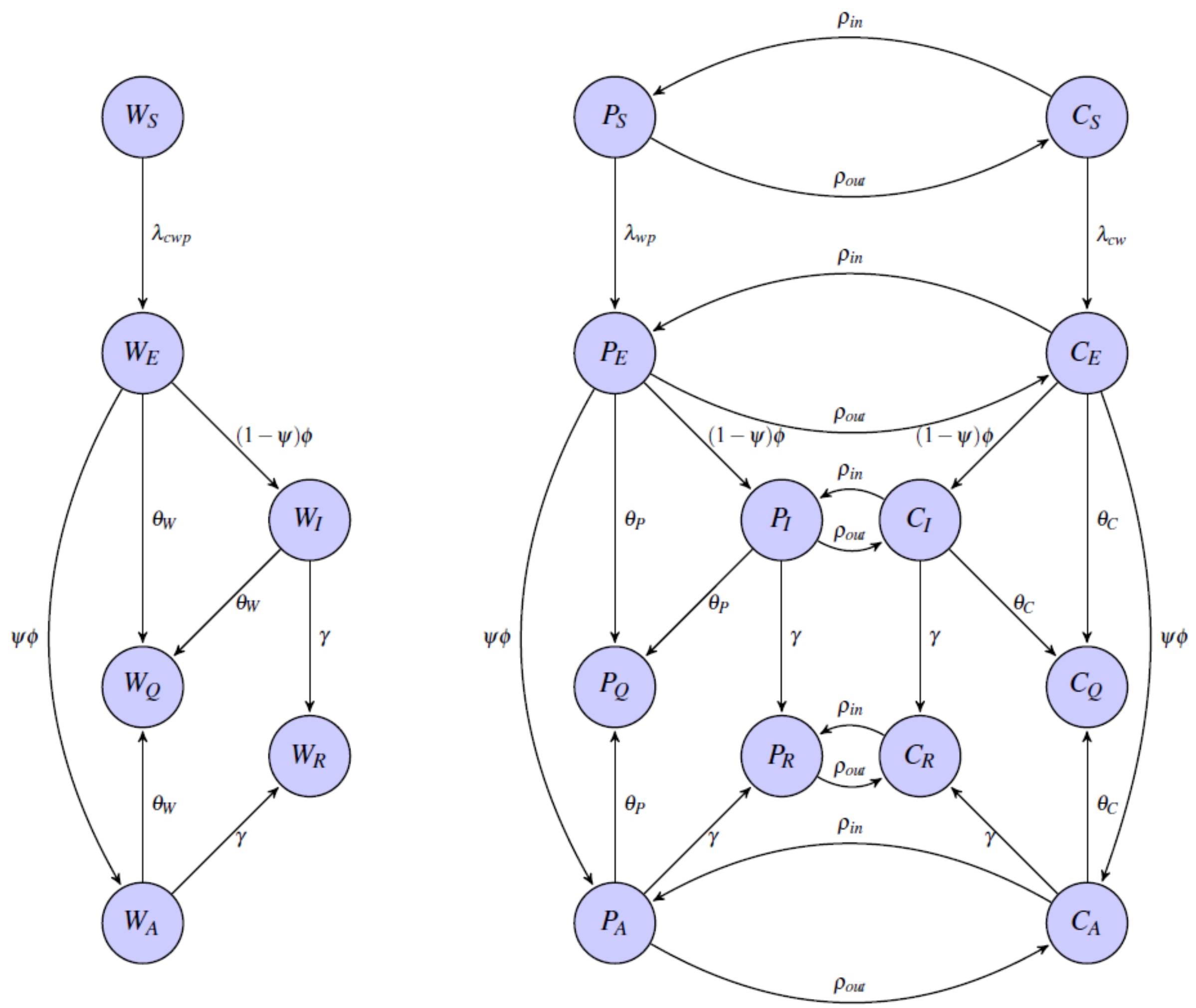


**Web Figure 1.** Compartmental Diagram for COVID-19 infection and incarceration

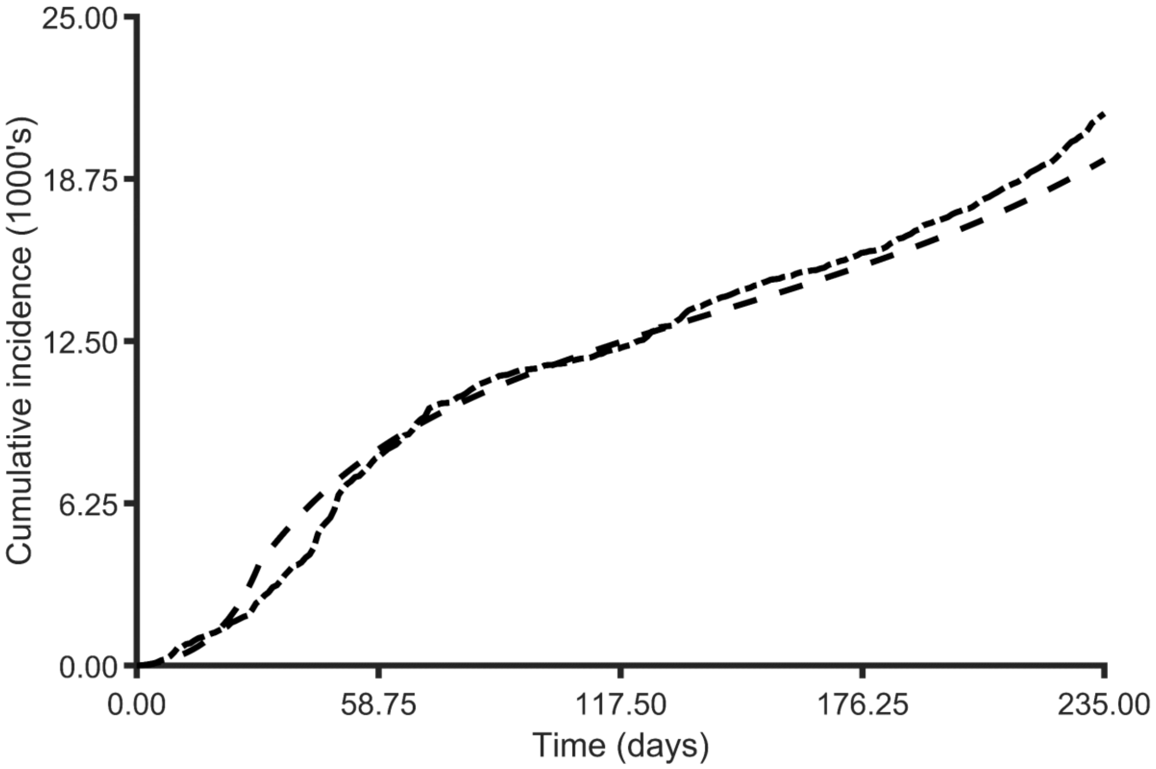


**Web Figure 2.** Fit of model to COVID-19 data. Estimated symptomatic and asymptomatic cumulative incidence of COVID-19 in Albany New York (dash dot line) and estimated cumulative incidence based on fitted ODE model (dashed line).

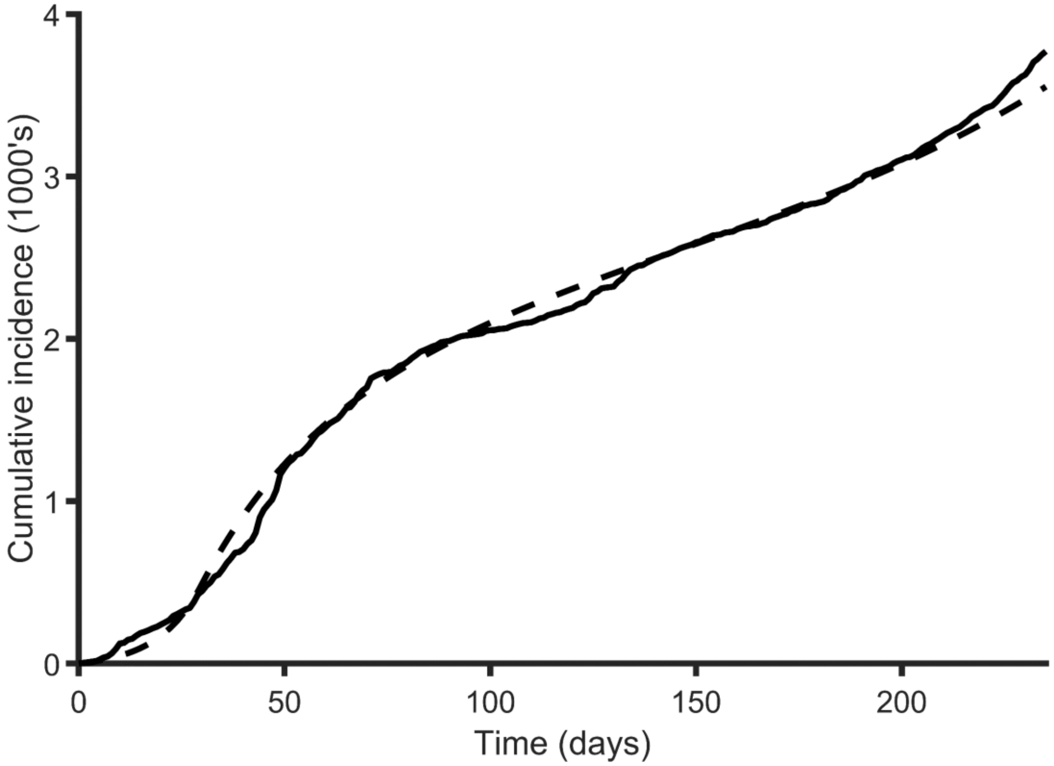


**Web Figure 3.** Fit of model to quarantine data. Cumulative positive tests of COVID-19 in Albany New York (dash dot line) and estimated quarantined infected individuals based on fitted ODE model (dashed line).

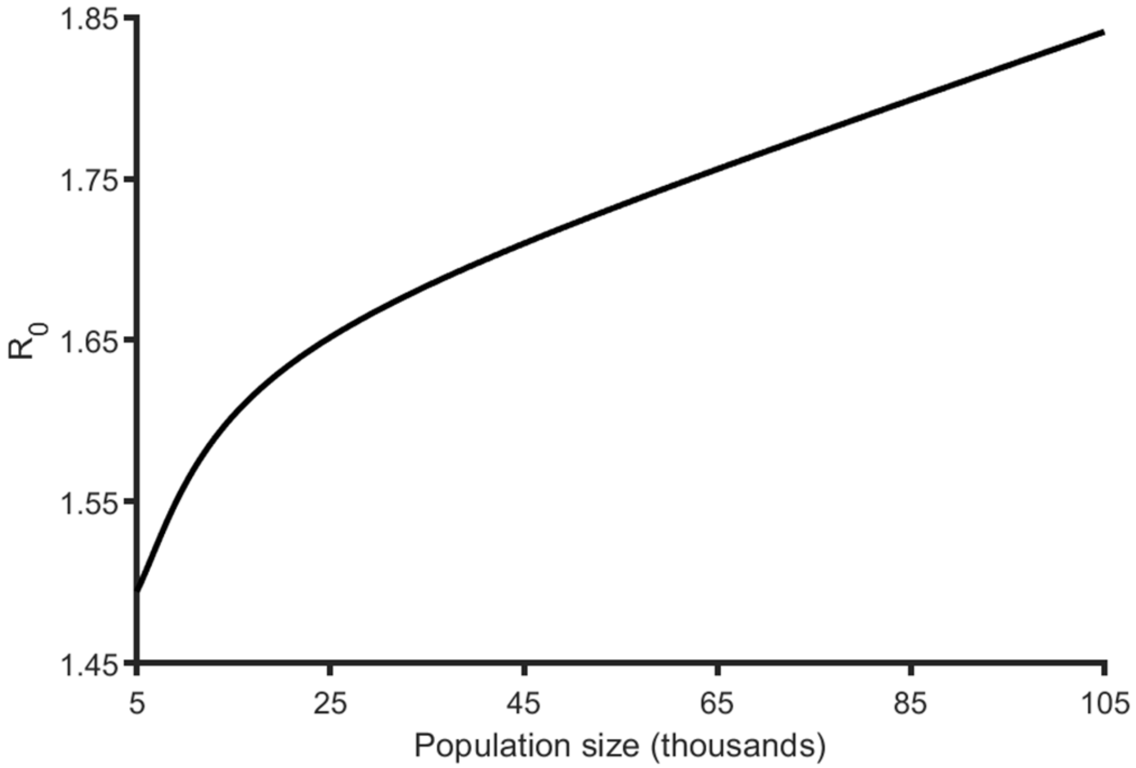


Web Figure 4. Basic reproductive number for the local community.

Web Table 1. COVID-19 associated transition rates for incarcerated persons

| Transition | Rate |
|---|---|
| $P_s \rightarrow P_s - 1$<br>$P_E \rightarrow P_E + 1$ | $\lambda_{CW} P_S \Delta t$ |
| $P_E \rightarrow P_E - 1$<br>$P_A \rightarrow P_A + 1$ | $\psi \phi P_E \Delta t$ |
| $P_E \rightarrow P_E - 1$<br>$P_I \rightarrow P_I + 1$ | $(1 - \psi) \phi P_E \Delta t$ |
| $P_E \rightarrow P_E - 1$<br>$P_Q \rightarrow P_Q + 1$ | $q_c P_E \Delta t$ |
| $P_A \rightarrow P_A - 1$<br>$P_R \rightarrow P_R + 1$ | $\gamma P_A \Delta t$ |
| $P_A \rightarrow P_A - 1$<br>$P_Q \rightarrow P_Q + 1$ | $q_c P_A \Delta t$ |
| $P_I \rightarrow P_I - 1$<br>$P_R \rightarrow P_R + 1$ | $\gamma P_I \Delta t$ |
| $P_I \rightarrow P_I - 1$<br>$P_Q \rightarrow P_Q + 1$ | $q_c P_I \Delta t$ |

Web Table 2. COVID-19 associated transition rates for incarcerated workers

| Transition | Rate |
|---|---|
| $W_s \rightarrow W_s - 1$<br>$W_E \rightarrow W_E + 1$ | $\lambda_{CWP} W_S \Delta t$ |
| $W_E \rightarrow W_E - 1$<br>$W_A \rightarrow W_A + 1$ | $\psi \phi W_E \Delta t$ |
| $W_E \rightarrow W_E - 1$<br>$W_I \rightarrow W_I + 1$ | $(1 - \psi) \phi W_E \Delta t$ |
| $W_E \rightarrow W_E - 1$<br>$W_Q \rightarrow W_Q + 1$ | $q_c W_E \Delta t$ |
| $W_A \rightarrow W_A - 1$<br>$W_R \rightarrow W_R + 1$ | $\gamma W_A \Delta t$ |
| $W_A \rightarrow W_A - 1$<br>$W_Q \rightarrow W_Q + 1$ | $q_c W_A \Delta t$ |
| $W_I \rightarrow W_I - 1$<br>$W_R \rightarrow W_R + 1$ | $\gamma W_I \Delta t$ |
| $W_I \rightarrow W_I - 1$<br>$W_Q \rightarrow W_Q + 1$ | $q_c W_I \Delta t$ |

Web Table 3. COVID-19 associated transition rates for the general community

| Transition | Rate |
|---|---|
| $C_s \rightarrow C_s - 1$<br>$C_E \rightarrow C_E + 1$ | $\lambda_{CW} C_S \Delta t$ |
| $C_E \rightarrow C_E - 1$<br>$C_A \rightarrow C_A + 1$ | $\psi \phi C_E \Delta t$ |
| $C_E \rightarrow C_E - 1$<br>$C_I \rightarrow C_I + 1$ | $(1 - \psi) \phi C_E \Delta t$ |
| $C_E \rightarrow C_E - 1$<br>$C_Q \rightarrow C_Q + 1$ | $q_c C_E \Delta t$ |
| $C_A \rightarrow C_A - 1$<br>$C_R \rightarrow C_R + 1$ | $\gamma C_A \Delta t$ |
| $C_A \rightarrow C_A - 1$<br>$C_Q \rightarrow C_Q + 1$ | $q_c C_A \Delta t$ |
| $C_I \rightarrow C_I - 1$<br>$C_R \rightarrow C_R + 1$ | $\gamma C_I \Delta t$ |
| $C_I \rightarrow C_I - 1$<br>$C_Q \rightarrow C_Q + 1$ | $q_c C_I \Delta t$ |

Web Table 4. Transition rates between the general community and incarceration facilities

| Transition | Rate |
|---|---|
| $C_s \rightarrow C_s - 1$<br>$P_s \rightarrow P_s + 1$ | $\rho_{in} C_s$ |
| $C_E \rightarrow C_s - 1$<br>$P_E \rightarrow P_E + 1$ | $\rho_{in} C_E$ |
| $C_I \rightarrow C_I - 1$<br>$P_I \rightarrow P_I + 1$ | $\rho_{in} C_I$ |
| $C_A \rightarrow C_A - 1$<br>$P_A \rightarrow P_A + 1$ | $\rho_{in} C_A$ |
| $C_R \rightarrow C_R - 1$<br>$P_R \rightarrow P_R + 1$ | $\rho_{in} C_R$ |
| $C_Q \rightarrow C_Q - 1$<br>$P_Q \rightarrow P_Q + 1$ | $\rho_{in} C_Q$ |
| $C_s \rightarrow C_s + 1$<br>$P_s \rightarrow P_s - 1$ | $\rho_{out} P_s$ |
| $C_E \rightarrow C_s + 1$<br>$P_E \rightarrow P_E - 1$ | $\rho_{out} P_E$ |
| $C_I \rightarrow C_I + 1$<br>$P_I \rightarrow P_I - 1$ | $\rho_{out} P_I$ |
| $C_A \rightarrow C_A + 1$<br>$P_A \rightarrow P_A - 1$ | $\rho_{out} P_A$ |
| $C_R \rightarrow C_R + 1$<br>$P_R \rightarrow P_R - 1$ | $\rho_{out} P_R$ |
| $C_Q \rightarrow C_Q + 1$<br>$P_Q \rightarrow P_Q - 1$ | $\rho_{out} P_Q$ |
